# The Causal Effects of Blood Iron and Copper on Lipid Metabolism Disease: Evidence from Phenome-wide Mendelian Randomization Study

**DOI:** 10.1101/2020.09.17.20196956

**Authors:** Jingqi Zhou, Chang Liu, Michael Francis, Yitang Sun, Moon-Suhn Ryu, Arthur Grider, Kaixiong Ye

## Abstract

**Background:** Blood levels of iron and copper, even within their normal ranges, have been associated with a wide range of clinical outcomes. Available epidemiological evidence on blood iron and copper association with potential clinical effects, such as lipid metabolism disorder, is inconsistent and scarce. This study aims to examine and disentangle the causal clinical effects of iron and copper.

**Methods:** Genetic instruments for the blood level of iron and copper were curated from existing genome-wide association studies. Candidate clinical outcomes were identified based on a phenome-wide association study (PheWAS) between these genetic instruments and multiple phenotypes in 310,999 unrelated individuals of European ancestry from the UK Biobank. All signals passing stringent correction for multiple testing were followed by Mendelian randomization (MR) analyses, with replication in independent data sources where possible.

**Results:** Genetically predicted higher blood levels of iron and copper are both associated with lower risks of iron deficiency anemia (OR = 0.75, 95% CI:0.67-0.85, *p* =1.90×10^−06^ for iron; OR= 0.88, 95% CI: 0.78-0.98, *p* =0.03 for cooper), lipid metabolism disorders and its two subcategories, hyperlipidemia (OR = 0.90, 95% CI:0.85-0.96, *p* =6.44×10^−04^ for iron; OR = 0.92, 95% CI:0.87-0.98, *p* = 5.51×10^−03^ for cooper) and hypercholesterolemia (OR = 0.90, 95% CI:0.84-0.95, *p* = 5.34×10^−04^ for iron; OR = 0.93, 95% CI:0.89–0.99, *p* =0.02 for cooper). Consistently, they are also associated with lower blood levels of total cholesterol and low-density lipoprotein cholesterol. Multiple sensitivity tests were applied to assess the pleiotropy and stability of the estimation, and consistent evidence across different approaches was obtained. Additionally, unique clinical effects of each blood mineral were also pinpointed, and sex-stratified MR analysis further revealed the clinical implication of iron and copper exhibits some degree of sex differences.

**Conclusions:** Our comparative PheWAS-MR study of iron and copper comprehensively characterized their shared and unique clinical effects, highlighting their novel causal roles in hyperlipidemia and hypercholesterolemia. Given the modifiable and variable nature of blood mineral status, these findings warrant further investigation.

## Introduction

Iron (Fe) and copper (Cu) are two essential mineral nutrients for human health through its vital role in enzymatic reactions and cellular energy metabolism ^1, 2^. Physiological processes that maintain these minerals homeostasis are overwhelmed by dietary loss, inappropriate iron absorption, inflammation, infection, liver disease, and disordered erythropoiesis. Disturbance to the systemic iron or copper homeostasis, including deficiency, excess, and even fluctuations in the normal ranges, could have clinical implications^3^. Iron deficiency, the most widespread micronutrient deficiency worldwide, is well-established to cause anemia, while iron overload can lead to chronic liver disease, cirrhosis, and hepatocellular carcinoma ^4, 5^. Copper deficiency is responsible for the lethal Menkes syndrome, featured by developmental delay and intellectual disability, while copper overload results in Wilson’s disease, characterized by hepatic cirrhosis and neurological deficits ^6^. Furthermore, copper is essential for iron absorption, and copper deficiency impairs iron metabolism. The multiple interactions of copper and iron involving both the positive and negative aspects by which associated with various of disease ^7^. For instance, epidemiological studies have reported elevated blood levels of iron and copper are associated with a higher risk of type 2 diabetes anemia and osteoarthritis (OA) ^8, 9^. Higher iron and copper have been associated with a reduced risk of hyperlipidemia and an improved blood lipid profile ^10, 11^. However, conflicting associative patterns were also reported ^12^. Given the crucial role of minerals, studying the health risks of iron and copper has important clinical guiding significance. Due to the limited data available and the fact that most studies were observational, it is uncertain whether the associations are causal and not biased by reverse causality or residual confounding. Moreover, the causality of iron and copper effect on overall health has not been clearly established.

Mendelian randomization (MR), a complementary approach to epidemiological observations, utilizes genetic variants as instrumental variables to approximate the lifetime status of an exposure (e.g., the blood level of a mineral) and evaluates its causal effect on a clinical outcome. The random allocation of alleles at conception and the natural direction of causality from genetic variants to phenotypes protect MR estimates from confounding and reverse causality ^13, 14^. MR studies have been performed to evaluate the causality of specific mineral-outcome pairs, such as iron on stroke, coronary artery disease (CAD), Parkinson’s disease, gout, and inflammatory bowel disease ^15, 16^; copper on ischemic heart disease ^17^. Compared to these hypothesis-driven MR studies with an obvious bias towards cardiovascular diseases, phenome-wide association study coupled with MR (PheWAS-MR) enables an unbiased and hypothesis-free scan through a wide range of phenotypes (i.e., phenome) and prioritized candidate clinical outcomes for MR causal inferences. Most importantly, few existing studies simultaneously examine multiple blood minerals at a phenome-wide scale to disentangle their confounded clinical effects ^18^.

In this study, we systematically evaluated and compared the causal clinical effects of iron and copper. Genetic instruments for the blood level of iron and copper were curated from existing genome-wide association studies (GWAS) with the largest sample sizes. Candidate clinical outcomes were identified based on a phenome-wide association study between these genetic instruments and multiple disease outcomes from the UK Biobank. All signals passing stringent correction for multiple testing were followed by Mendelian randomization (MR) analyses, with replication in independent data sources where possible. Additionally, we examined the causal association between iron or copper levels and a subset of lipid profiles, which are essential biomarkers for lipid metabolism disease, in the secondary analysis with an independent dataset.

## Methods

The guideline of Strengthening the Reporting of Observational Studies in Epidemiology (STROBE) (S1 Checklist) was followed in this study report. The UK Biobank project was approved by the North West Multi-Centre Research Ethics Committee and appropriate informed consent was obtained from participants. Data used in the project was accessed through an approved application to UK Biobank. Only a subset of unrelated White British individuals with high-quality genotype data (N = 310,999) were included in this study in order to minimize the complication of population structure. The study design of PheWAS-MR, replication, and sensitivity analysis was prespecified. A schematic of the overall study design is shown in Fig 1.

**Fig 1:**
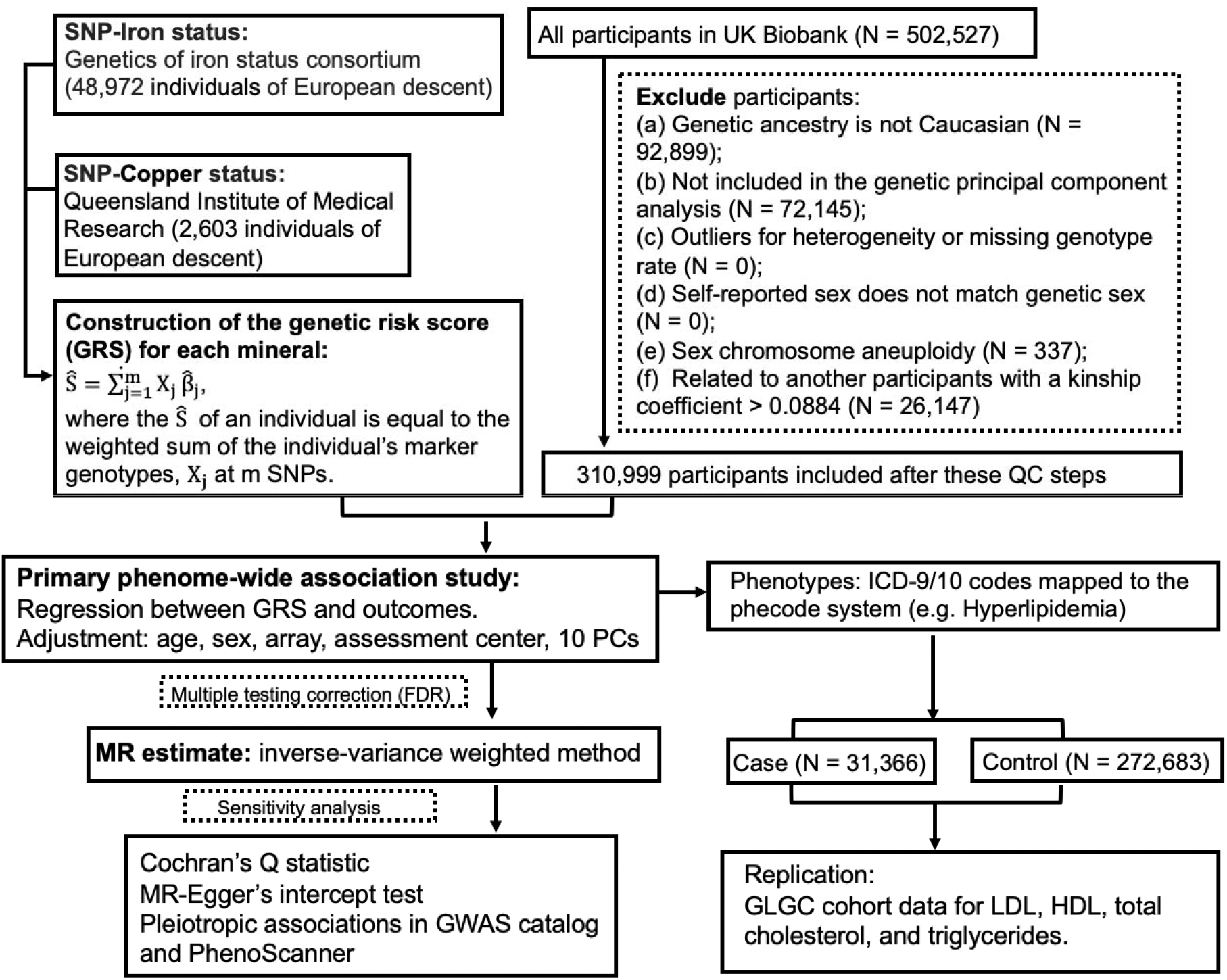
A flow chart of the study design.

### Genetic instruments for blood iron and copper

Genetic instrumental variables for the blood levels of iron and copper were selected from previous GWAS (Table 1). Based on a meta-analysis of 48,972 subjects of European descent performed by the Genetics of Iron Status consortium ^19^, three single nucleotide polymorphisms (SNPs) were selected as genetic instruments for systemic iron status. All three of them are consistently associated with four iron status biomarkers, including serum iron, ferritin, transferrin, and transferrin saturation. Their effects on serum iron were used in MR analysis. They are very strong genetic instruments, with F-statistics ranging from 445 to 796 and collectively explaining approximately 3.8% of the variation in serum iron. They have been previously used in MR analysis of iron status ^20^. Genetic variants associated with erythrocyte copper levels were identified in an Australian cohort (N = 2603) ^21^. Two SNPs were selected for erythrocyte copper, which accounted for 4.60% of the phenotypic variance. For each blood mineral, a weighted polygenic risk score (GRS) was constructed for each UK Biobank participant by summing over selected SNPs weighted by their effect sizes.

**Table 1:** SNPs used as genetic instruments for blood minerals in the PheWAS and MR analyses.

| SNP | Effect allele | Baseline allele | Chr | Closest gene | % variance explained | F-statistic | EAF | Beta<sup>a</sup> | SE | P |
| --- | --- | --- | --- | --- | --- | --- | --- | --- | --- | --- |
| 3 SNPs for Fe from Benyamin et al. (N<sup>b</sup> = 48,972) |  |  |  |  |  |  |  |  |  |  |
| rs1800562 | A | G | 6 | HFE | 1.30 | 645 | 0.067 | 0.328 | 0.016 | 2.72×10<sup>-97</sup> |
| rs1799945 | G | C | 6 | HFE | 0.90 | 445 | 0.15 | 0.189 | 0.010 | 1.10×10<sup>-81</sup> |
| rs855791 | G | A | 22 | TMPRSS6 | 1.60 | 796 | 0.554 | 0.181 | 0.007 | 1.32×10<sup>-139</sup> |
| 2 SNPs for Cu from Evans et al. (N<sup>b</sup> = 2,603) |  |  |  |  |  |  |  |  |  |  |
| rs1175550 | G | A | 1 | SMIM1 | 1.45 | 38 | 0.23 | 0.198 | 0.032 | 5.03×10<sup>-10</sup> |
| rs2769264 | G | T | 1 | SELENBP1 | 3.15 | 85 | 0.18 | 0.313 | 0.034 | 2.63×10<sup>-20</sup> |

SNP single nucleotide polymorphism, Chr: chromosome, EAF: effect allele frequency, SE: standard error,
a. The beta coefficient of mineral increasing allele on concentrations of serum iron (in $\mu\text{mol/L}$ ), erythrocyte copper (in $\text{mmol/L}$ ).
b. The sample size of the GWAS or meta-analysis from which the genetic variants were selected.

### Study population

The UK Biobank is a population-based prospective cohort established to study genetic and environmental determinants of human diseases. More than 500,000 individuals aged 40-69 years were recruited between 2006 and 2010, all of whom gave written consent and underwent baseline measurements. Participants donated blood samples for genotyping and biomarker analysis. Moreover, UK Biobank participants consented access to their electronic health records. Only participants fulfilling the following criteria were included in our analysis: genetic ancestry of Caucasian, included in genetic principal component analysis, not outliers for heterogeneity and missing genotype rate, no sex chromosome aneuploidy, no high degree of genetic kinship (i.e., ten or more third-degree relatives identified), and self-reported sex matching genetic sex. Additionally, for remaining pairs of relatives (kinship coefficient > 0.0884), a minimum number of participants were removed so that all remaining participants are unrelated. Analysis was restricted to participants of Caucasian descent in order to maintain consistency with the European samples used to obtain genetic instruments for blood mineral status. A total of 310,999 unrelated individuals were included in our final analysis.

### Phenome wide association study

Three phenotypic datasets in the UK Biobank, including inpatient hospital records, cancer registry, and death registry, were included in this analysis. First, we used the International Classification of Diseases (ICD) versions 9 and 10 to identify cases in the hospital episode statistics, including both incident and prevalent cases, but not self-reported diagnoses. As ICD-9 / ICD-10 codes are not organized as independent phenotypes, we mapped them to the phecode system of distinct diseases or traits ^22^. The mapping process also automatically excludes patients with similar or potentially overlapping diseases from the corresponding control group. The online phecode map is accessible via the link https://phewascatalog.org/phecodes_icd10. We restricted analysis to phecodes with sufficient numbers of cases to ensure more than 80% statistical power in subsequent MR analyses. The number of cases and controls in analyses of phenotypes for each blood mineral is shown in S1 Table.

For each mineral, case-control groups were generated for each phecode, and logistic regression was performed, separately, for each instrument SNP, adjusting for age, sex, genotyping array, assessment center, and the first 10 genetic principal components. Moreover, we explored the association between the weighted GRS of each blood mineral and the corresponding phecodes with logistic regression. Given that phenotypes investigated are not totally independent even in the phecode system, we applied the false discovery rate (FDR) method with a cutoff of 0.05 to correct for multiple testing.

### MR analyses

To assess whether blood mineral levels have causal effects on the candidate clinical outcomes identified by the PheWAS, we further conducted two-sample MR analyses, where the instrument-mineral and instrument-outcome associations were estimated in two different samples. In this study, the instrument-mineral effect was estimated in previous GWAS, while the instrument-outcome effect was estimated in the above-mentioned PheWAS. To estimate the causal effect of each mineral on a phenotype, for each genetic instrument, we calculated the ratio of its effect on the phenotype to its effect on the blood mineral. An overall effect estimate across multiple genetic instruments was achieved with three methods, the inverse variance-weighted (IVW) method, weighted median (WM) method, and MR-Egger ^23, 24^. Effect estimates, measured as OR of the outcome, were normalized to 1 standard deviation (SD) increment in each blood mineral. Additionally, sex-stratified IVW MR estimates were obtained with sex-specific instrument-outcome effects from genetically males and females, respectively. Mineral-outcome pairs exhibiting evidence of horizontal pleiotropy were excluded in this stratified analysis.

To validate some mineral-outcome relationships that we discovered based on UK Biobank, we performed replication analysis based on different cohorts, which were accessed through the MR-Base ^25^. Specifically, for the four blood lipids, including total cholesterol (TC), high-density lipoprotein cholesterol (HDL), low-density lipoprotein cholesterol (LDL), and triglyceride, we extracted the instrument-outcome summary statistics from a previous GWAS by the Global Lipids Genetics Consortium (GLGC) ^26^.

To further evaluate if the effects of two minerals on a shared clinical outcome are independent of each other, we adopted a multivariable MR framework ^27^, in which the effects of multiple related risk factors can be estimated simultaneously. For instance, to test if the effects of iron and copper on a shared clinical outcome (e.g., hyperlipidemia) are independent, we included all the genetic instruments for iron and copper in the analysis. The effect sizes of instrument SNPs on serum iron and erythrocyte copper were extracted from previous GWAS ^19, 21^, while their effects on the shared clinical outcome were calculated in our curated UK Biobank dataset. We modelled the SNP effects on the shared outcome (β_shared_) against the SNP effects on iron (β_Fe_), adjusted for the effects on copper (β_Cu_), using a weighted linear regression model (β_shared_∼β_Fe_ + β_Cu_), where the weights were defined by inverse standard errors of β_shared_.

### Sensitivity analyses

Several sensitivity analyses were performed to detect and correct for the presence of pleiotropy in the causal estimates. We began with the fixed-effect IVW MR approach, and calculated Cochran’s Q statistic for heterogeneity, with which a p-value ≤ 0.05 indicates the presence of pleiotropy across genetic instruments. Heterogeneity across SNP-outcome associations will lead to bias in the fixed-effect IVW estimate. If substantial heterogeneity exists, we proceeded to a random-effect IVW approach, which allows instruments to exhibit balanced horizontal pleiotropy.

Because the IVW estimate is essentially a weighted average of the Wald ratios obtained from each SNP, if any of the SNPs exhibit horizontal pleiotropy, then the causal effect estimate is liable to be biased. We further performed WM MR ^24^ and MR-Egger analyses ^28^. The WM analysis calculates the median of an empirical distribution of MR association estimates weighted by their precisions. It provides consistent estimates when more than half of the instruments are valid ^24^. The MR-Egger method provides an intercept test, with a non-zero intercept indicating the presence of unbalanced horizontal pleiotropy. It also provides an unbiased estimate of the causal effect, taking into account pleiotropy. When there was no heterogeneity across instruments, a fixed-effect model was used to estimate the causal effect. Otherwise a random-effect model was used ^28^.

Besides the above-mentioned sensitivity analyses that leverage information from multiple genetic instruments for one blood mineral, for each individual genetic instrument, we also evaluated their pleiotropic effects by searching the GWAS catalog and PhenoScanner V2 ^29^, to identify any secondary phenotypes associated (*p* < 5 × 10^−8^) with them or their proxies (r^2^ > 0.8). All analyses were conducted using the MendelianRandomisation ^30^ and TwoSampleMR ^25^ packages, and the R programming language.

## Results

After quality controls on the full UK Biobank dataset, a subset of 310,999 unrelated individuals of European descent were included in the PheWAS analysis. Main demographic characteristics of the study population and basic statistics of all genetic instruments are summarized in S2 Table. All instrument SNPs satisfied the Hardy-Weinberg equilibrium test (*p* > 0.05). The association between the weighted GRS of each blood mineral and common confounding factors (i.e., age, sex, genotyping array, assessment center, and the genetic principal components) are presented in S3 Table. All these factors were controlled for in the PheWAS analysis.

PheWAS analysis was limited to phenotypes (i.e., phecodes) that had enough cases to ensure more than 80% statistical power in the subsequent MR analyses. Among them, 12 clinical outcomes, representing 6 disease categories, reached statistical significance at the 5% FDR threshold. Next, we performed MR analyses to examine the possible causal links between each of the significant outcomes identified in PheWAS. Mineral-outcome pairs exhibit consistent MR evidence for causal effects, without suggestions of heterogeneity and horizontal pleiotropy across genetic instruments based on Cochran’s Q statistic and MR-Egger’s intercept test were provided in test (S4 Tables).

### Shared and unique causal clinical effects of blood iron and copper

Iron and copper share some of causally associated clinical outcomes, including iron deficiency anemia, disorders of lipoid metabolism, hypercholesterolemia, and hyperlipidemia (Fig 2). IVW MR analyses provided significant evidence for causal effects of both higher blood iron and copper on reduced risks of iron deficiency anemia (OR per SD increase in serum iron = 0.75, 95%CI: 0.67-0.85, *p* =1.81×10^−6^; OR per SD increase in erythrocyte copper = 0.88, 95% CI: 0.79-0.99, *p* = 0.032) and disorders of lipoid metabolism (OR per SD increase in serum iron = 0.90, 95% CI: 0.85-0.96, *p* =6.61×10^−4^; OR per SD increase in erythrocyte copper = 0.92, 95% CI: 0.87-0.98, *p* = 4.94×10^−3^;), and its two subtypes, hyperlipidemia (OR = 0.90, CI: 0.85-0.96, *p* = 6.44×10^−4^; OR = 0.92, CI: 0.87-0.98, *p* = 5.51×10^−3^) and hypercholesterolemia (OR = 0.90, CI: 0.84-0.95, p = 5.34×10^−4^; OR = 0.93, CI: 0.88–0.99, *p* = 0.022). Specifically, unique causal clinical effects were identified for iron and copper. Per SD increase in serum iron is associated with enhanced risk of varicose veins (OR = 1.28, CI: 1.15-1.42, *p* = 4.34×10^−6^) and acquired foot deformities (OR = 1.21, CI: 1.09-1.35, *p* = 4.95×10^−4^), but with reduced risk of other anemia (OR = 0.72; CI: 0.65-0.79; *p* = 1.12×10^−11^). Per SD increment in erythrocyte copper is associated with increased risk of osteoarthrosis (OA) (OR = 1.07, CI: 1.02-1.13, *p* = 0.010).

**Fig 2.**
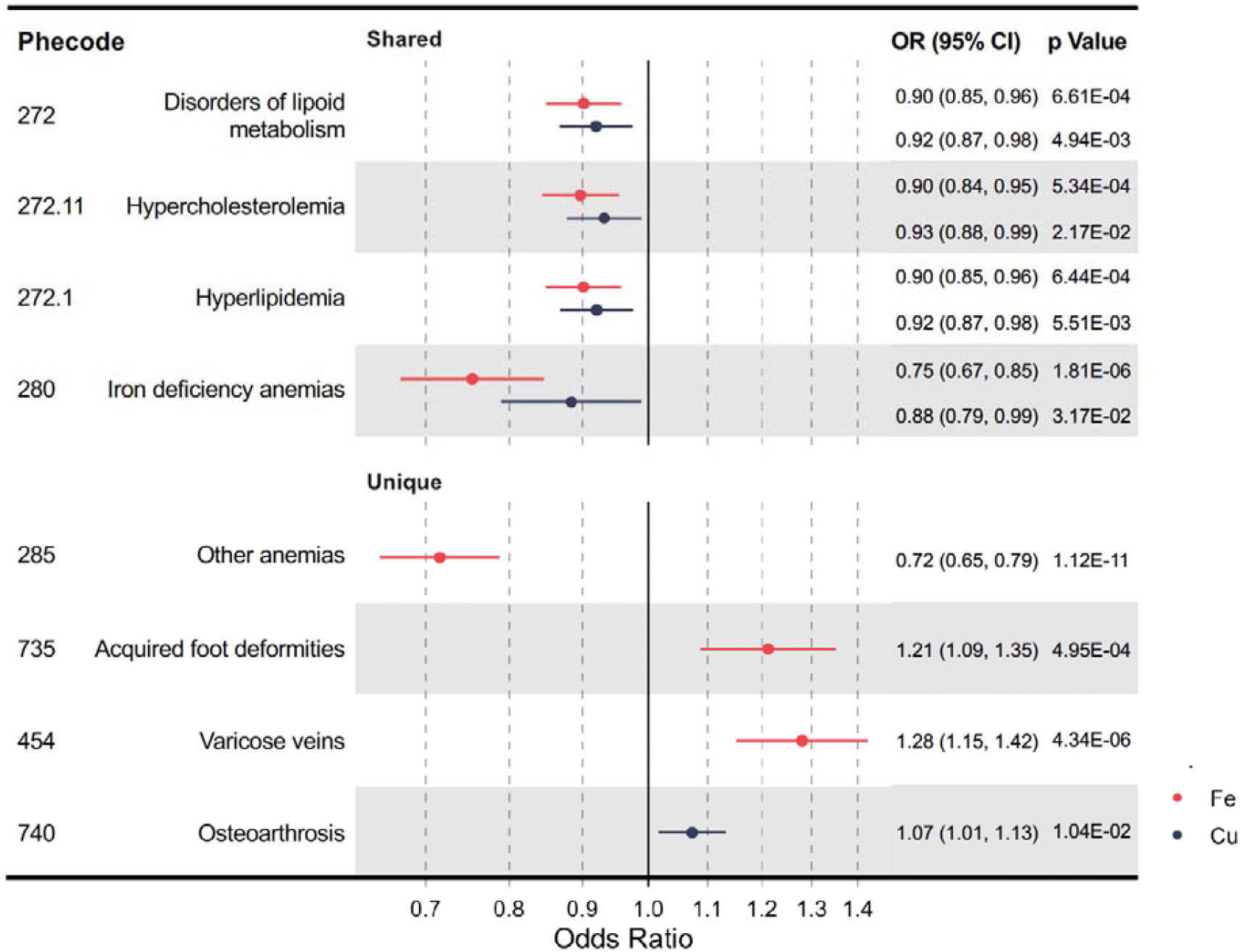
A forest plot showing significant mineral-outcome associations based on MR analysis. The causal estimates are from IVW MR and have no indications of pleiotropy. The odds ratios (ORs) with their 95% confidence intervals (CIs) are scaled to 1-SD increase in blood iron or copper level. Complete MR results are provided in S4 Tables.

### Causality of both Iron and Cooper on Lipid Metabolism Trait

We performed further investigation into the shared clinical effects of serum iron and erythrocyte copper on lipid metabolism disorders, by first evaluating if they are independent of each other and then by examining their causal effects on blood biomarkers of lipid metabolism, including TC, HDL, LDL, and triglyceride. First, we adopted a multivariable MR framework to estimate the direct effects of serum iron and erythrocyte copper on their shared clinical outcomes, by conditioning on the effects of the other mineral. We found that the causal effects of iron and copper are independent of each other and do not show much differences compared to those of univariate MR (S5 Table). Based on the shared but independent effects of iron and coper on lipid metabolism disorders, we further hypothesized that their genetically predicted levels are associated with blood biomarkers of lipid metabolism. We tested this hypothesis with the UK Biobank data with exclusion of participants under statin medication, and then in an independent replication cohort from the Global Lipids Genetics Consortium (GLGC) accessed through the MR-base database (Table 2). In both cohorts, genetically predicted higher levels of serum iron is associated with decreased levels of LDL (WM OR = 0.91, CI: 0.89-0.94, *p* = 5.27×10^−11^ in UKBB) and TC (WM OR = 0.91, CI: 0.88-0.94, *p* = 8.99×10^−8^ in UKBB), but elevated levels of triglycerides (WM OR = 1.05, CI: 1.02-1.07, *p* = 4.2×10^−5^ in UKBB). No significant effects were found for HDL. For erythrocyte copper, while no significant effects were found in UK Biobank, analysis in the GCLC cohort showed that per SD increase in copper is associated with reduced levels of LDL (OR = 0.95, CI: 0.92-0.99, *p* = 0.011 in GCLC) and TC (OR = 0.96, CI: 0.92-0.99, *p* =0.02 in GCLC). Overall, we found evidence for the shared effects of iron and copper on reducing blood levels of LDL and TC.

**Table 2:**
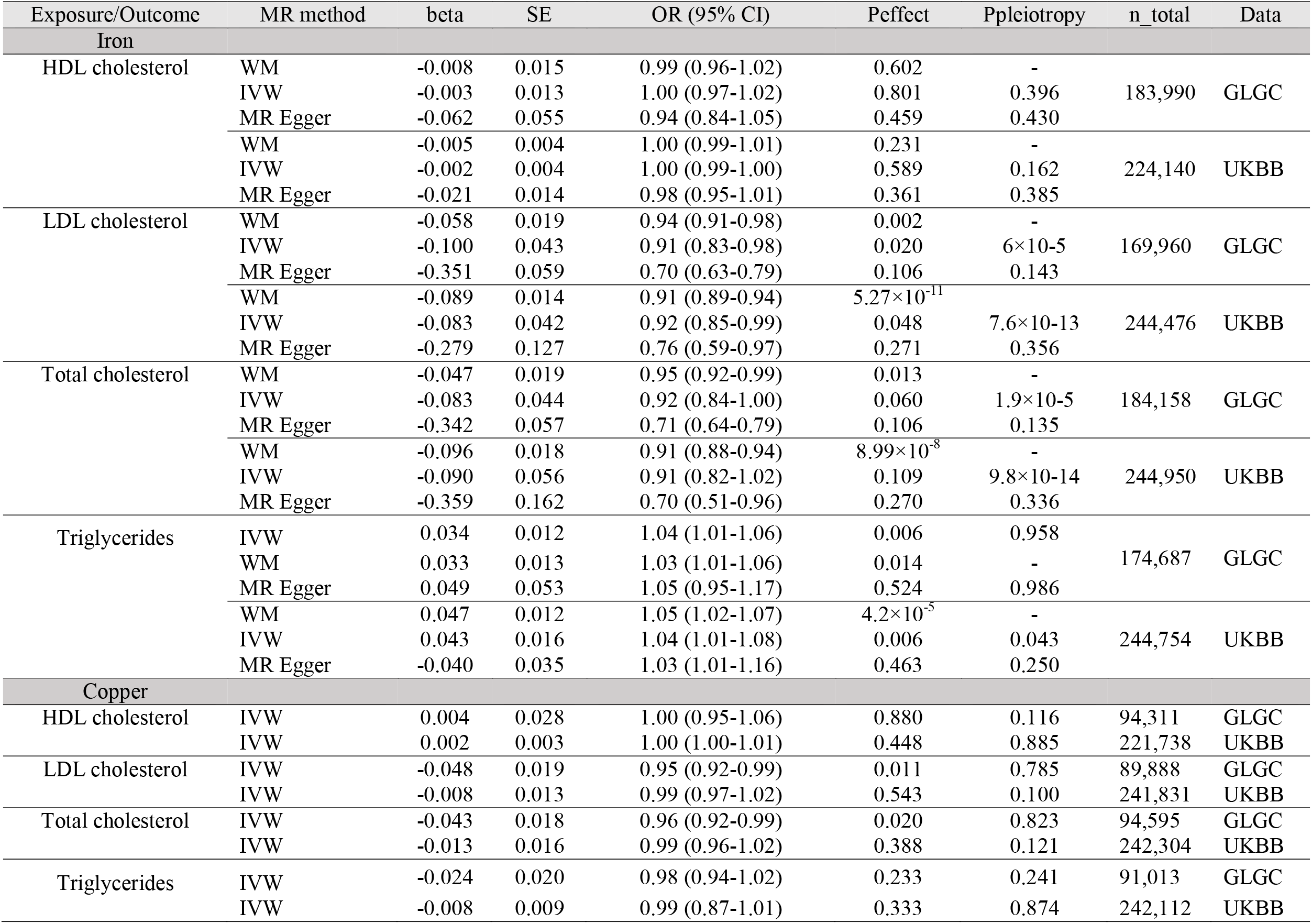
MR analyses of iron and copper on blood lipids in UK Biobank and GLGC

| Exposure/Outcome | MR method | beta | SE | OR (95% CI) | Peffect | Ppleiotropy | n_total | Data |
| --- | --- | --- | --- | --- | --- | --- | --- | --- |
| Iron |  |  |  |  |  |  |  |  |
| HDL cholesterol | WM | -0.008 | 0.015 | 0.99 (0.96-1.02) | 0.602 | - | 183,990 | GLGC |
|  | IVW | -0.003 | 0.013 | 1.00 (0.97-1.02) | 0.801 | 0.396 |  |  |
|  | MR Egger | -0.062 | 0.055 | 0.94 (0.84-1.05) | 0.459 | 0.430 |  |  |
|  | WM | -0.005 | 0.004 | 1.00 (0.99-1.01) | 0.231 | - | 224,140 | UKBB |
|  | IVW | -0.002 | 0.004 | 1.00 (0.99-1.00) | 0.589 | 0.162 |  |  |
|  | MR Egger | -0.021 | 0.014 | 0.98 (0.95-1.01) | 0.361 | 0.385 |  |  |
| LDL cholesterol | WM | -0.058 | 0.019 | 0.94 (0.91-0.98) | 0.002 | - | 169,960 | GLGC |
|  | IVW | -0.100 | 0.043 | 0.91 (0.83-0.98) | 0.020 | 6×10-5 |  |  |
|  | MR Egger | -0.351 | 0.059 | 0.70 (0.63-0.79) | 0.106 | 0.143 |  |  |
|  | WM | -0.089 | 0.014 | 0.91 (0.89-0.94) | 5.27×10<sup>-11</sup> | - | 244,476 | UKBB |
|  | IVW | -0.083 | 0.042 | 0.92 (0.85-0.99) | 0.048 | 7.6×10-13 |  |  |
|  | MR Egger | -0.279 | 0.127 | 0.76 (0.59-0.97) | 0.271 | 0.356 |  |  |
| Total cholesterol | WM | -0.047 | 0.019 | 0.95 (0.92-0.99) | 0.013 | - | 184,158 | GLGC |
|  | IVW | -0.083 | 0.044 | 0.92 (0.84-1.00) | 0.060 | 1.9×10-5 |  |  |
|  | MR Egger | -0.342 | 0.057 | 0.71 (0.64-0.79) | 0.106 | 0.135 |  |  |
|  | WM | -0.096 | 0.018 | 0.91 (0.88-0.94) | 8.99×10<sup>-8</sup> | - | 244,950 | UKBB |
|  | IVW | -0.090 | 0.056 | 0.91 (0.82-1.02) | 0.109 | 9.8×10-14 |  |  |
|  | MR Egger | -0.359 | 0.162 | 0.70 (0.51-0.96) | 0.270 | 0.336 |  |  |
| Triglycerides | IVW | 0.034 | 0.012 | 1.04 (1.01-1.06) | 0.006 | 0.958 | 174,687 | GLGC |
|  | WM | 0.033 | 0.013 | 1.03 (1.01-1.06) | 0.014 | - |  |  |
|  | MR Egger | 0.049 | 0.053 | 1.05 (0.95-1.17) | 0.524 | 0.986 |  |  |
|  | WM | 0.047 | 0.012 | 1.05 (1.02-1.07) | 4.2×10<sup>-5</sup> | - | 244,754 | UKBB |
|  | IVW | 0.043 | 0.016 | 1.04 (1.01-1.08) | 0.006 | 0.043 |  |  |
|  | MR Egger | -0.040 | 0.035 | 1.03 (1.01-1.16) | 0.463 | 0.250 |  |  |
| Copper |  |  |  |  |  |  |  |  |
| HDL cholesterol | IVW | 0.004 | 0.028 | 1.00 (0.95-1.06) | 0.880 | 0.116 | 94,311 | GLGC |
|  | IVW | 0.002 | 0.003 | 1.00 (1.00-1.01) | 0.448 | 0.885 | 221,738 | UKBB |
| LDL cholesterol | IVW | -0.048 | 0.019 | 0.95 (0.92-0.99) | 0.011 | 0.785 | 89,888 | GLGC |
|  | IVW | -0.008 | 0.013 | 0.99 (0.97-1.02) | 0.543 | 0.100 | 241,831 | UKBB |
| Total cholesterol | IVW | -0.043 | 0.018 | 0.96 (0.92-0.99) | 0.020 | 0.823 | 94,595 | GLGC |
|  | IVW | -0.013 | 0.016 | 0.99 (0.96-1.02) | 0.388 | 0.121 | 242,304 | UKBB |
| Triglycerides | IVW | -0.024 | 0.020 | 0.98 (0.94-1.02) | 0.233 | 0.241 | 91,013 | GLGC |
|  | IVW | -0.008 | 0.009 | 0.99 (0.87-1.01) | 0.333 | 0.874 | 242,112 | UKBB |

### Interpretation of potential pleiotropy in PheWAS-MR results

To detect and correct for any possible pleiotropic effect of multiple instruments, we then conducted the different sets of sensitivity analysis. All causal clinical effects of iron and copper presented above in combined analysis, except LDL and TC, do not have evidence for heterogeneity or horizontal pleiotropy across genetic instruments, as indicated by Cochran’s Q statistic and MR-Egger’s intercept test (S4 Tables). For some relationships, causal estimates from another methods, WM MR and MR-Egger, are broadly consistent with those from IVW MR, although with wider confidence intervals as expected from their lower statistical powers ^31^. For instance, the causal estimates of serum iron on disorders of lipoid metabolism were consistent in the IVW MR (OR = 0.90, CI: 0.85-0.96, *p* = 6.61×10^−4^) and WM MR (OR= 0.91, 95% CI:0.85-0.97, *p* = 6.25×10^−3^), and there was no evidence of pleiotropy (*p* ≥ 0.05 for all related tests).

Besides sensitivity analyses that leverage information from multiple genetic instruments for one blood mineral, for each individual instrument SNP, we also evaluated their pleiotropic effects by searching the GWAS catalog and PhenoScanner V2 ^29^. The HFE rs1800562 variant is associated with low-density lipoprotein cholesterol ^26^, and this SNP were identified as pleiotropic with potential confounders. Similar magnitudes of associations were observed when these variants were excluded from the MR analyses. We additionally performed a sensitivity analysis of sex-stratified IVW MR to identify sex-specific causal clinical effects of these minerals. We identified mineral-outcome pairs exhibiting sex-specific causal relationships (Fig 3, Supplementary Tables 6). That is, their MR estimates are significant only in one sex group, but not in the other or the sex-combined sample. Iron is associated with higher risks of myeloproliferative diseases (OR =2.20, CI: 1.37-3.53, *p* =1.14×10^−3^) in females and chronic liver disease (OR =1.47, CI: 1.07-2.03, *p* =0.017) in males. Specifically, copper increases the risk of diabetes mellitus (OR = 1.20, CI: 1.02-1.40, *p* = 0.027) and its subcategory, T2D (OR = 1.19, CI: 1.00-1.40, *p* = 0.044) in female. But serum iron and copper have no sex-specific effects on lipid metabolism disease.

**Fig 3.**
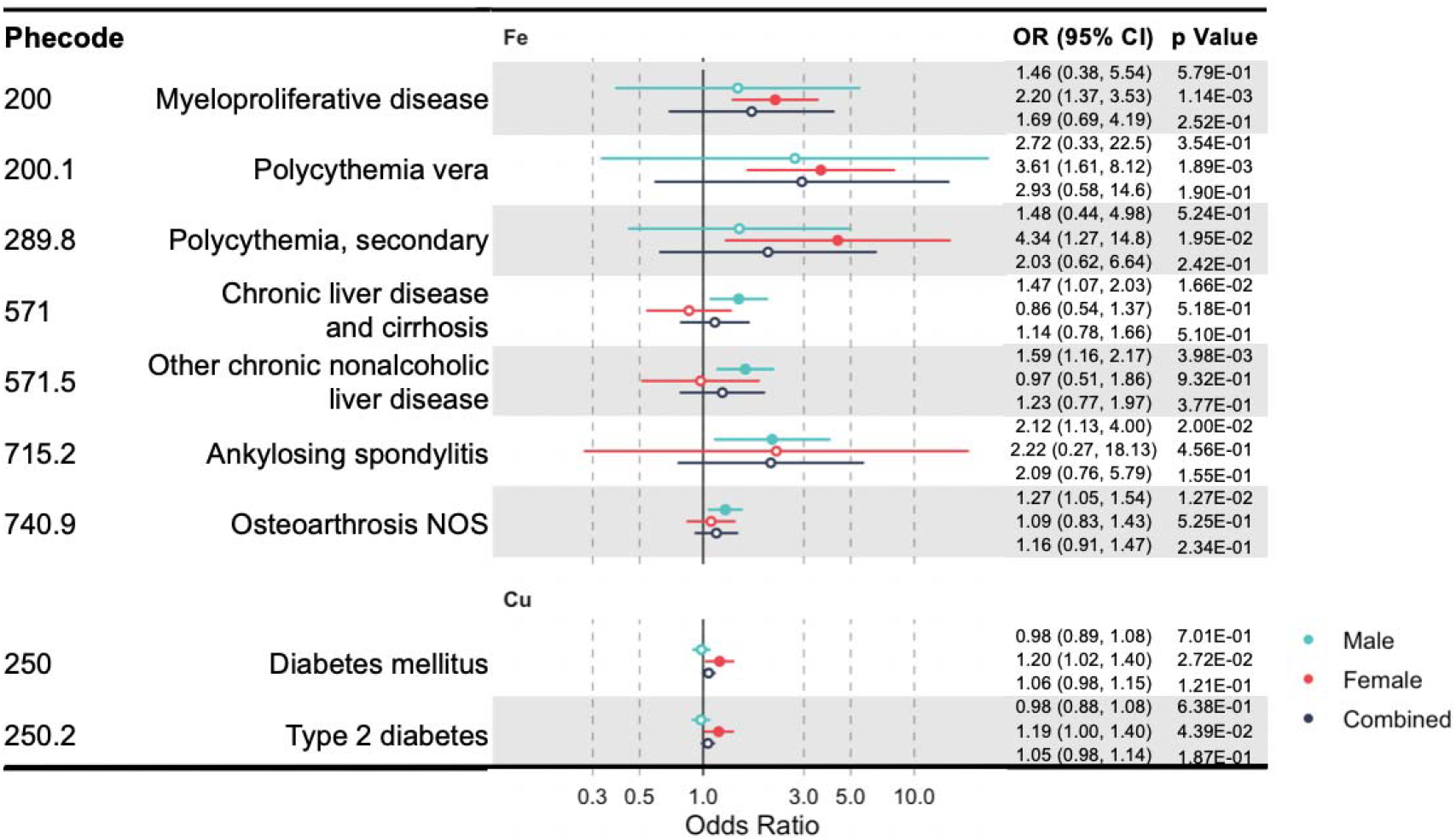
A forest plot showing significant mineral-outcome associations based on sex-stratified MR analysis. Results in male, female, and the combined analyses are shown. The causal estimates are from IVW MR and have no indications of pleiotropy. The odds ratios (ORs) with their 95% confidence intervals (CIs) are scaled to 1-SD increase in blood mineral level. Complete MR results are provided in S6 Tables.

## Discussion

Our study adopted a phenome-wide approach to systematically evaluate and compare the clinical effects of blood iron and copper minerals. This is the first PheWAS-MR study for copper, and the first to perform systematic comparison across iron and copper. Our findings highlighted shared clinical effects, specifically the protective effects of iron and copper on lipid metabolism disorders and iron deficiency anemia. Some causal mineral-outcome relationships identified in this study have been well known and supported by existing mechanistic studies, while others are novel, awaiting further confirmation and future mechanistic exploration.

Most notably, we found that genetically predicted higher blood levels of iron and copper are both protective against lipid metabolism disorders and its two subcategories, hyperlipidemia and hypercholesterolemia. Consistently, they are also associated with lower blood TC and LDL. This is the first MR study to establish the causal protective effect of copper on lipid metabolism disorders. The interplays among lipid, iron, and copper metabolism have been recognized, but they are complicated and far away from being fully elucidated. Copper deficiency have long been linked to increased risks of hyperlipidemia and cardiovascular diseases in both humans and animal models ^32^, while copper supplementation in patients of hyperlipidemia was shown to improve the blood lipid profile ^33^. Epidemiological findings, however, are conflicting, with some suggesting negative associations of serum copper with both TC and LDL ^34^, and others showing positive associations ^35^. The relationship between iron status and blood lipids is similarly convoluted. On the one hand, some epidemiological studies found that serum iron is lower in hyperlipidemic patients ^36^, and that serum ferritin is negatively associated with LDL ^11^. Very likely related, elevated iron status is negatively associated with coronary artery disease in both observational and MR studies ^37, 38^. Moreover, studies on rats showed that iron deficiency upregulates lipogenic genes but downregulates apolipoprotein H and genes involved in the mitochondrial beta-oxidation, resulting in increased circulating lipids ^39^. On the other hand, anemic and/or iron-deficient patients and animal models tend to have lower blood TC and LDL ^40^, while serum ferritin has often been found to be positively associated with an unhealthy lipid profile (i.e., elevated TC, LDL, and triglyceride, but decreased HDL) ^41^. To gain mechanistic insights into how iron regulates lipid metabolism, we performed bioinformatic search for iron responsive elements in genes involved in cholesterol metabolism and fatty acid degradation ^42^. Multiple potential iron-responsive genes were identified, including LIPH and LDLRAP1 (S1-S2 Figs). Our study highlights iron and copper as risk factors of dyslipidemia and calls for future studies into their physiological mechanism.

Well-known mineral-outcome relationships with established physiological mechanisms provide support for the power and validity of our study. Iron is an integral component of the oxygen-carrying hemoglobin and is required for the process of erythropoiesis ^4^. In keeping with this, we found that genetically predicted higher level of serum iron is protective against anemia, while in women it increases the risks of myeloproliferative disease, polycythemia vera, and secondary polycythemia. Myeloproliferative diseases are bone marrow and blood disorders featured by abnormal hematopoiesis, while its most common subtype, polycythemia vera is characterized by erythrocytosis (i.e., excessive red blood cell production) ^43^. Very interestingly, iron deficiency without anemia is present in virtually all patients of polycythemia vera ^44^. Our findings suggest that iron deficiency is the result, rather than the cause, of erythrocytosis and reaffirm the current practice of not using iron supplement to treat patients of polycythemia vera. At the other extreme of systemic status, iron overload can cause tissue damage and its excessive accumulation in liver leads to cirrhosis and hepatocellular carcinoma ^3^. Consistently, our study revealed that elevated serum iron in men is positively associated with risks of chronic liver disease and cirrhosis, and one of its subtypes, other chronic nonalcoholic liver disease. For copper, its shared protective effect with iron on iron deficiency anemia also has relatively well-established mechanistic basis. First, the efflux of iron from enterocyte, hepatocyte, and macrophage into the blood circulation requires the action of two copper-dependent ferroxidases (i.e., hephaestin and ceruloplasmin), which oxidize ferrous iron into ferric iron so that it could be loaded onto transferrin. Besides enabling iron transport, copper is also required for hemoglobin biosynthesis, probably by assisting iron import into or utilization in mitochondria ^45^.

Our study revealed the novel causal roles of elevated blood iron in increased risk of varicose vein. Varicose vein, a common venous disease of the lower extremity, is characterized by incompetent valves, reflux, and venous wall dilation. Its etiological process involves the hydrostatic-pressure-induced activation of matrix degrading enzymes and inflammatory cascade ^46^. Iron overload and its causal HFE genetic variations have been associated with varicose vein ^47^.

Mechanistically, iron overload induces oxidative stress and hyperactivation of matrix degrading enzymes ^48^. Other significant mineral-outcome relationships are also worth attention. For iron, our finding of its status increases the risk of glossitis (i.e., tongue inflammation) has been reported in a previous MR study ^49^. However, it is fairly well established that patients of atrophic glossitis frequently suffer from deficiencies of nutrients, including iron, and corresponding nutrient supplementation is able to resolve oral symptoms ^50^. How to reconcile these findings should be a subject of future studies. Our findings open many new venues of research to elucidate the roles of blood iron and copper in these clinical conditions.

Our study has strengths and limitations. This is the first comparative PheWAS-MR of iron and copper, based on a large prospective cohort, to simultaneously evaluate their shared and unique causal clinical effects. Our approach is unbiased and hypothesis-free. The MR approach reduces the bias from confounding and reserve causality. We utilized strong genetic instruments and applied multiple sensitivity analyses to detect the presence of pleiotropy. Only MR estimates with no indications of pleiotropy were reported. For selected outcomes (i.e., blood lipids), we replicated the results with an independent dataset. The sex-stratified analysis enabled the identification of sex-specific relationships. Our results confirmed some previous MR findings while made novel discoveries, most of which are supported by existing epidemiological and mechanistic studies. Our study also has multiple limitations. Despite a large sample size (N = 310,999) of the overall cohort, the case numbers of specific outcomes are still small, limiting our statistical power for rare diseases and those with modest effects. Moreover, since our genetic instruments only proxy blood mineral levels in the normal ranges, some clinical effects of extreme levels, such as deficiency and overload, are likely missed. Another related issue is that MR estimation assumes a linear relationship between the exposure and the outcome, and as a result, non-linear effects might have been missed. Given the well-known threshold effects of mineral deficiency and overload, future studies with a non-linear model will likely reveal more clinical effects. Genetic instruments approximate the average effects over the life course, while the physiological relevance of a blood mineral could vary by life stages and is not captured by our study. Finally, UK Biobank is a middle- and old-aged cohort and we restricted analysis to those of European descent, future studies in cohorts of younger ages or other ethnic backgrounds are needed to confirm our findings and to search for more clinical consequences.

## Conclusions

In conclusion, our comparative PheWAS-MR study of iron and copper comprehensively characterized their shared and unique clinical effects. These known and novel mineral-outcome relationships provide profound insights into the heath impacts of normally varying blood minerals and into the etiologies of some clinical conditions, specifically hyperlipidemia and hypercholesterolemia. Our findings also emphasize the possibility and importance of managing blood minerals, probably through dietary adjustment, in the prevention and management of these medical conditions.

## Data Availability

Genetic and phenotypic data from UK Biobank could be accessed following an application and approval process. Summary statistics generated in this project are provided in the supplementary files.

## Acknowledgments

This study was conducted using the UK Biobank Resource under project 48818. The authors would like to thank all UK Biobank participants. We also want to thank other members of the Ye lab for stimulating discussions.

## Abbreviations

CAD: coronary artery disease
FDR: false discovery rate
GLGC: Global Lipids Genetics Consortium
ICD: International Classification of Diseases
IVW: inverse variance-weighted
GRS: polygenic risk score
GWAS: genome-wide association study
HDL: high-density lipoprotein cholesterol
LDL: low-density lipoprotein cholesterol
MR: mendelian randomization
OR: odds ratio
OA: Osteoarthrosis
PheWAS: phenome-wide association study
SNP: single nucleotide polymorphism
SD: standard deviation
T2D: type 2 diabetes
TC: total cholesterol
WM: weighted median

## Author Contributions

Conceptualization: Jingqi Zhou, Kaixiong Ye.

Data Curation: Jingqi Zhou, Chang Liu, Michael Francis.

Formal Analysis: Jingqi Zhou, Chang Liu.

Funding Acquisition: Moon-Suhn Ryu, Arthur Grider, Kaixiong Ye.

Investigation: Jingqi Zhou, Chang Liu, Arthur Grider, Kaixiong Ye.

Methodology: Jingqi Zhou, Kaixiong Ye.

Project Administration: Jingqi Zhou, Kaixiong Ye.

Resources: Moon-Suhn Ryu, Arthur Grider, Kaixiong Ye.

Software: Jingqi Zhou, Chang Liu, Michael Francis, Yitang Sun.

Supervision: Kaixiong Ye.

Validation: Michael Francis, Yitang Sun, Kaixiong Ye.

Visualization: Jingqi Zhou, Chang Liu.

Writing – Original Draft Preparation: Jingqi Zhou, Chang Liu.

Writing – Review & Editing: Jingqi Zhou, Chang Liu, Michael Francis, Yitang Sun, Moon-Suhn Ryu, Arthur Grider, Kaixiong Ye.

## Supporting Information

S1 Checklist: STROBE checklist. STROBE, Strengthening the Reporting of Observational Studies in Epidemiology.

S1 Fig: Genes in the KEGG pathway of cholesterol metabolism predicted to carry iron response elements. Predictions with high, medium, and low confidence are shown in yellow, blue, and orange, respectively.

S2 Fig: Genes in the KEGG pathway of fatty acid degradation predicted to carry iron response elements. Predictions with high, medium, and low confidence are shown in yellow, blue, and orange, respectively.

S1 Table: The number of phenotypes and cases considered in each disease category.

S2 Table: Descriptive characteristics of the UK Biobank participants and the SNPs included in PheWAS analyses.

S3 Table: The association between the weighted GRS of each blood mineral and common confounding factors.

S4 Table: Lists of significant mineral-outcome relationships without indications of pleiotropy. S5 Table: Multivarate IVW MR results for iron and copper.

S6 Table: Lists of significant mineral-outcome relationships without indications of pleiotropy based on sex-stratified MR analysis.

